# Race and ethnicity-related differences in neuroimaging markers of neurodegeneration and cerebrovascular disease in middle and older age

**DOI:** 10.1101/2021.01.15.21249876

**Authors:** Indira C. Turney, Patrick J. Lao, Miguel Arce Rentería, Kay Igwe, Joncarlos Berroa, Andres Rivera, Andrea Benavides, Clarissa Morales, Nicole Schupf, Richard Mayeux, Jose Gutierrez, Jennifer J. Manly, Adam M. Brickman

**Affiliations:** Taub Institute for Research on Alzheimer’s Disease and the Aging Brain, College of Physicians and Surgeons, Columbia University, New York, NY 10032, USA; Gertrude H. Sergievsky Center, College of Physicians and Surgeons, Columbia University, New York, NY 10032, USA; Department of Neurology, College of Physicians and Surgeons, Columbia University, New York, NY 10032, USA

## Abstract

**INTRODUCTION:** Numerous neuroimaging studies demonstrated racial and ethnic disparities in brain health at older ages. It remains unclear whether racial and ethnic disparities increase with aging and whether they are already apparent in midlife.

**METHODS:** We investigated differences in MRI markers of aging and cerebrovascular disease in 969 participants from the Washington Heights-Inwood Columbia Aging Project (WHICAP; mean age: 75 years) and 496 participants from the Offspring study (mean age: 55 years) across race and ethnicity (white, Black, Latinx).

**RESULTS:** Older whites had greater cortical thickness compared with Latinxs, who also had greater thickness than Blacks. Cortical thickness was similar across race in the middle-aged cohort. Regarding white matter hyperintensity (WMH) volume, Blacks had disproportionately greater WMH volume compared to both whites and Latinxs at older ages. Racial disparities are already apparent in midlife, where Blacks have disproportionately greater WMH than whites.

**Discussion:** These findings suggest that racial disparities in WMH volume are already apparent in midlife.

## 1. Background

Stressful lifetime events are more prevalent among Blacks and Latinx than among whites^1–4^ and are associated with cardiovascular disease^5,6^ and inflammation^7,8^. Blacks are more likely than whites to experience chronic stress in the form of material hardship, interpersonal discrimination, structural discrimination, ambient stressors, segregated housing, and personal danger^9^. The “Weathering Hypothesis” suggests that repeated exposure to stress, dangerous environments, and general social disadvantage contributes to accelerated wear and tear, leading to a faster rate of aging in Blacks and Latinxs compared with whites^10^. The Weathering Hypothesis has been proposed as a framework to understand differential health outcomes across race and ethnicity groups in late life^10,11^, including with respect to brain aging^12^. Indeed, previous studies reported racial and ethnic differences in neuroimaging markers of aging and neurodegeneration in older adults. For example, DeCarli and colleagues^13^ showed that older Blacks had smaller hippocampal volume than whites, particularly among those with a family history of dementia. Similarly, we previously reported increased markers of cerebrovascular disease, including white matter hyperintensities (WMH)^14^, in Blacks and Latins Latinxs compared with white older adults.

It is unclear at what point in the adult lifespan racial and ethnic disparities in neuroimaging markers of brain aging emerge. While previous studies showed differences across race and ethnicity groups in late life, correlations of age with markers of brain aging generally did not differ across groups^13,14^, suggesting that race- or ethnicity-related differences emerge earlier in life. The purpose of the current study was to examine differences in neuroimaging markers of brain aging, including regional cortical thickness and WMH volume, across race and ethnicity groups in middle age and late life. We hypothesized that differences between race/ethnicity groups in late life would already be apparent in middle-aged adults.

## 2. Methods

### 2.1. Participants

We selected participants from the Washington Heights-Inwood Columbia Aging Project (WHICAP) and Offspring cohorts, two community-based studies of cognitive aging and dementia in residents of northern Manhattan, New York. Full descriptions of study procedures have been published previously^15^. Briefly, WHICAP participants were recruited in three waves, beginning in 1992, 1999, and 2009. Magnetic resonance imaging (MRI) was introduced into the WHICAP study in 2005^16^ with 1.5T scanning; current analyses include participants recruited from the 2009 cohort where 1016 participants received structural scans with 3T MRI beginning in 2011^17^. Offspring participants are adult children of WHICAP participants, age 28 or older, fluent in English or Spanish, willing to donate a sample of blood, and willing to be followed longitudinally. To date, we have enrolled 1,469 Offspring, 549 of whom received MRI scanning. MRI participants were a random subset of the overall study cohort, intending to scan one participant per family. Only participants with complete MRI and covariates data were considered for the analyses, resulting in a sample size of 1465. Of the 1465 total participants included in our analysis, there were only 193 family units, including siblings and parent-child dyads (104 parents with 1 offspring, 60 parents with 2 offspring, 14 parents with 3 offspring, 10 parents with 4 offspring, 2 parents with 5 offspring, 2 parents with 6 offspring, and 1 parent with 8 offspring).

The overall subset of participants across both studies included in the present study (n=1465) met the following inclusion criteria: (1) underwent at least partial neuropsychological evaluation at the time of their MRI, (2) self-reported their race/ethnicity as white (non-Latinx; hereafter referred to as white), African-American (non-Latinx; hereafter referred to as Black) or Latinx (any race), and (3) had complete data on quantitative MRI variables of interest (i.e., WMH volume and cortical thickness). All participants gave written informed consent and received compensation for their participation. The Institutional Review Board at Columbia University Medical Center approved this study.

### 2.2. MRI acquisition and processing

Participants in WHICAP were scanned on a 3T Philips Intera scanner at Columbia University between 2011 and 2019. Parameters for the T1-weighted anatomical scans were: TR = 6.6 ms; TE =3.0 ms; flip angle = 8°; field of view (FOV) = 256 ⨯ 200 ⨯ 165 mm^3^; in-plane resolution = 1 ⨯ 1 mm^2^; slices = 165. T2-weighted fluid attenuated inversion recovery (FLAIR; repetition time = 8000 ms, echo time = 337 ms, inversion time = 2400 ms, FOV = 240 ⨯ 240 ⨯ 180 matrix with 1 mm slice thickness) images were acquired in the axial orientation. Participants in Offspring were scanned on a 3T Siemens Prisma scanner at Columbia University between 2018 and 2020. Parameters for the T1-weighted anatomical scans were: TR = 2300 ms; TE =2.26 ms; flip angle = 8°; FOV = 256 ⨯ 256 ⨯ 192 mm^3^; in-plane resolution = 1 ⨯ 1 mm^2^; slices = 192. T2-weighted FLAIR (repetition time = 5000 ms, echo time = 387 ms, inversion time = 1800; FOV = 230 ⨯ 230 ⨯173 matrix with .90 mm slice thickness) images were acquired in the axial orientation.

Using previously described procedures^18–20^, whole-brain WMH volumes were quantified from T2-weighted fluid-attenuated inversion recovery (FLAIR) images with in-house developed software. Briefly, in WHICAP, images were brain extracted using FSL BET (Brain Extraction Tool)^21^, and a Gaussian curve was fit to map voxel intensity values. Voxels at 2.1 standard deviations above the image mean were labeled as WMH. In Offspring, T2-weighted FLAIR images were brain extracted using HD-Bet^22^, then corrected for intensity inhomogeneities using the ANTs N4 bias field correction algorithm^23^. High-pass filtering was then applied at the mode of distribution of the image voxel intensity values. We then fit a half Gaussian mixture model to the log-transformed histogram of the intensity values of each image. The Gaussian distribution that captured the highest intensity values were labeled as WMH. Any cluster of labeled voxels that included fewer than 5 voxels was removed from the mask. Across both studies, labeled images were also visually inspected and manually corrected if errors were detected (Figure 1a and 1b). Finally, a sum of the number of labeled voxels was then multiplied by voxel dimensions to yield a total volume in cm^3^.

**Figure 1.**
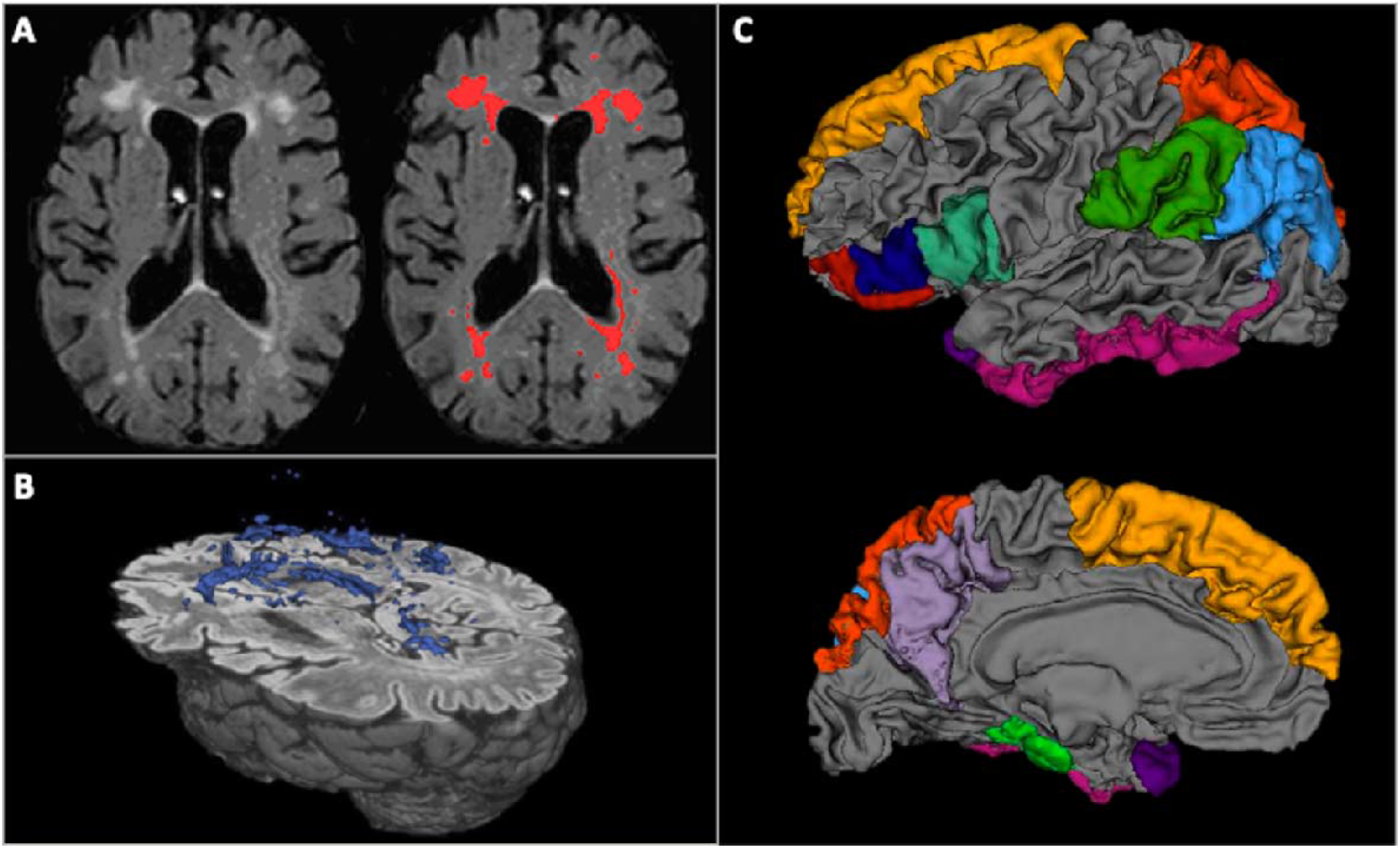
An axial slice displaying distribution of WMH unlabeled (A, left) with in-house developed software and WMH labeled (A, right). A three-dimensional rendering of the labeled WMH volume (B). A three-dimensional rendering of the anatomically segmented regions of interest included in the AD signature composite (C). Abbreviation: WMH, white matter hyperintensities.

Regional cortical thickness was quantified with FreeSurfer version 5.1 for WHICAP and 6.0 for Offspring (http://surfer.nmr.mgh.harvard.edu/) using T1-weighted images. A single “AD signature” measure was derived for each participant by averaging cortical thickness values across hemisphere in nine regions previously shown to best reflect AD neurodegeneration (Dickerson et al., 2009). These regions included: rostral medial temporal lobe (entorhinal cortex and parahippocampus), angular gyrus (inferior parietal lobe), inferior frontal lobe (pars opercularis, pars orbitalis, and pars triangularis), inferior temporal lobe (inferior temporal lobe), temporal pole (temporal pole), precuneus (precuneus), supramarginal gyrus (supramarginal gyrus), superior parietal lobe (superior parietal lobe), and superior frontal lobe (superior frontal lobe) (Figure 1c).

### 2.3. Statistical analysis

We used t-tests for continuous variables and chi-square for categorical variables to evaluate differences in baseline demographic characteristics between participants in each study, including age at scan, years of education, sex/gender, and race/ethnicity. To examine the pattern of race/ethnicity differences in brain health between studies, stratified by study (Offspring and WHICAP), we used ANOVAs to compare cortical thickness and WMH volumes across race and ethnicity groups. We examined these associations in stratified models in order to better understand cohort-specific effects, given variability between cohorts (e.g., age-span, life experiences, comorbidities). Covariates included age at scan and sex/gender. We compared effect sizes to determine the magnitude of differences across race/ethnicity groups between the two studies/cohorts.

Participants from the Offspring study are mainly middle-aged, however, there was some overlap in age across both studies. Sensitivity analysis examined these associations after excluding participants who were not more exclusively in the middle-aged or older age group (Offspring 25-65.99 years; WHICAP=66-98 years).

## 3. Results

### 3.1. Demographic Characteristics

Characteristics of WHICAP and Offspring participants are shown in Table 1. Compared with WHICAP, Offspring participants were younger at the time of scanning, had more years of education, and comprised a greater proportion of Latinx participants. The proportion of women was similar in the two studies.

**Table 1.** Demographics across study cohort

|  | **Offspring**<br>**(n = 496)** | **WHICAP**<br>**(n = 969)** | **Total**<br>**(n = 1465)** | **Statistic<sup>a</sup>** |
| --- | --- | --- | --- | --- |
| Age (years); M (SD) | 55.21 (10.66) | 75.18 (6.45) | 68.42 (12.46) | F=1985.26<sup>†</sup> |
| Education (years); M (SD) | 12.94 (3.46) | 12.20 (4.69) | 12.44 (4.34) | F=9.38<sup>^</sup> |
| Sex/Gender, n (%) | | | | $\chi^2=2.91$ |
| Women | 327 (65.3) | (589) 60.7 | 916 (62.3) | - |
| Race/Ethnicity, n (%) | | | | $\chi^2=131.85^{\dagger}$ |
| Non-Latinx White | 33 (6.6) | 243 (25.1) | 276 (18.8) | - |
| Non-Latinx Black | 118 (23.6) | 338 (34.8) | 456 (31.0) | - |
| Latinx | 350 (69.9) | 389 (40.1) | 739 (50.2) | - |

Abbreviations: M, mean; SD, standard deviation
<sup>a</sup> t-test or chi-square for demographics between study/age-groups
<sup>^</sup> $p < .01$ .
<sup>†</sup> $p < .001$ .

### 3.2. Imaging Results

The relationship of race/ethnicity with cortical thickness (Table 2) and WMH volume (Table 3) differed by study. In WHICAP, whites had greater cortical thickness than Latinxs, and Blacks and Latinxs had greater thickness than Blacks (main effect of race/ethnicity, F_2,969_=23.28, *p*<.001; η^2^=.05). In Offspring, cortical thickness was similar across race/ethnicity groups (main effect of race/ethnicity, F_2,496_=.50, *p=*.61; η^2^=.002) (Figure 2). In both studies, women had greater cortical thickness relative to men, and increased age was associated with lower cortical thickness.

**Table 2.** Parameter Estimates for cortical thickness stratified by study cohort

|  | B(SE) | p-values | 95% CI of Estimates |  |
| --- | --- | --- | --- | --- |
|  |  |  | Lower | Upper |
| *Offspring Study* |  |  |  |  |
| Age | -.003(.0004) | 1.14<sup>-13</sup> | -.004 | -.002 |
| Sex/Gender: Women | .03(.009) | .003 | .009 | .05 |
| Race/Ethnicity |  |  |  |  |
| Non-Latinx White | 0<sup>a</sup> | - | - | - |
| Non-Latinx Black | -.007(.02) | .733 | -.05 | .03 |
| Latinx | -.01(.02) | .434 | -.05 | .02 |
| *WHICAP Study* |  |  |  |  |
| Age | -.008(.001) | 4.37<sup>-39</sup> | -.009 | -.007 |
| Sex/Gender: Women | .03(.007) | .00002 | .02 | .05 |
| Race/Ethnicity |  |  |  |  |
| Non-Latinx White | 0<sup>a</sup> | - | - | - |
| Non-Latinx Black | -.06(.01) | .000 | -.08 | -.05 |
| Latinx | -.03(.009) | .002 | -.05 | -.01 |

Abbreviations: B, unstandardized parameter estimate; SE, standard error; CI, confidence interval; M, mean; SD, standard deviation
<sup>a</sup> reference group

**Table 3.** Parameter Estimates for WMH volume stratified by study cohort

|  | B(SE) | p-values | 95% CI of Estimates |  |
| --- | --- | --- | --- | --- |
|  |  |  | Lower | Upper |
| *Offspring Study* |  |  |  |  |
| Age | .008(.002) | 5.25E<sup>-7</sup> | .005 | .01 |
| Sex/Gender: Women | -.10(.04) | .003 | -.17 | -.04 |
| Race/Ethnicity |  |  |  |  |
| Non-Latinx White | 0<sup>a</sup> | - | - | - |
| Non-Latinx Black | .21(.08) | .010 | .05 | .37 |
| Latinx | .16(.08) | .033 | .01 | .31 |
| *WHICAP Study* |  |  |  |  |
| Age | .02(.003) | 9.37<sup>-11</sup> | .01 | .02 |
| Sex/Gender: Women | -.06(.03) | .078 | -.12 | .007 |
| Race/Ethnicity |  |  |  |  |
| Non-Latinx White | 0<sup>a</sup> | - | - | - |
| Non-Latinx Black | .10(.04) | .015 | .02 | .19 |
| Latinx | -.04(.04) | .389 | -.12 | .05 |

Abbreviations: WMH, white matter hyperintensity; B, unstandardized parameter estimate; SE, standard error; CI, confidence interval; M, mean; SD, standard deviation; ref, reference group
<sup>a</sup> reference group

**Figure 2.**
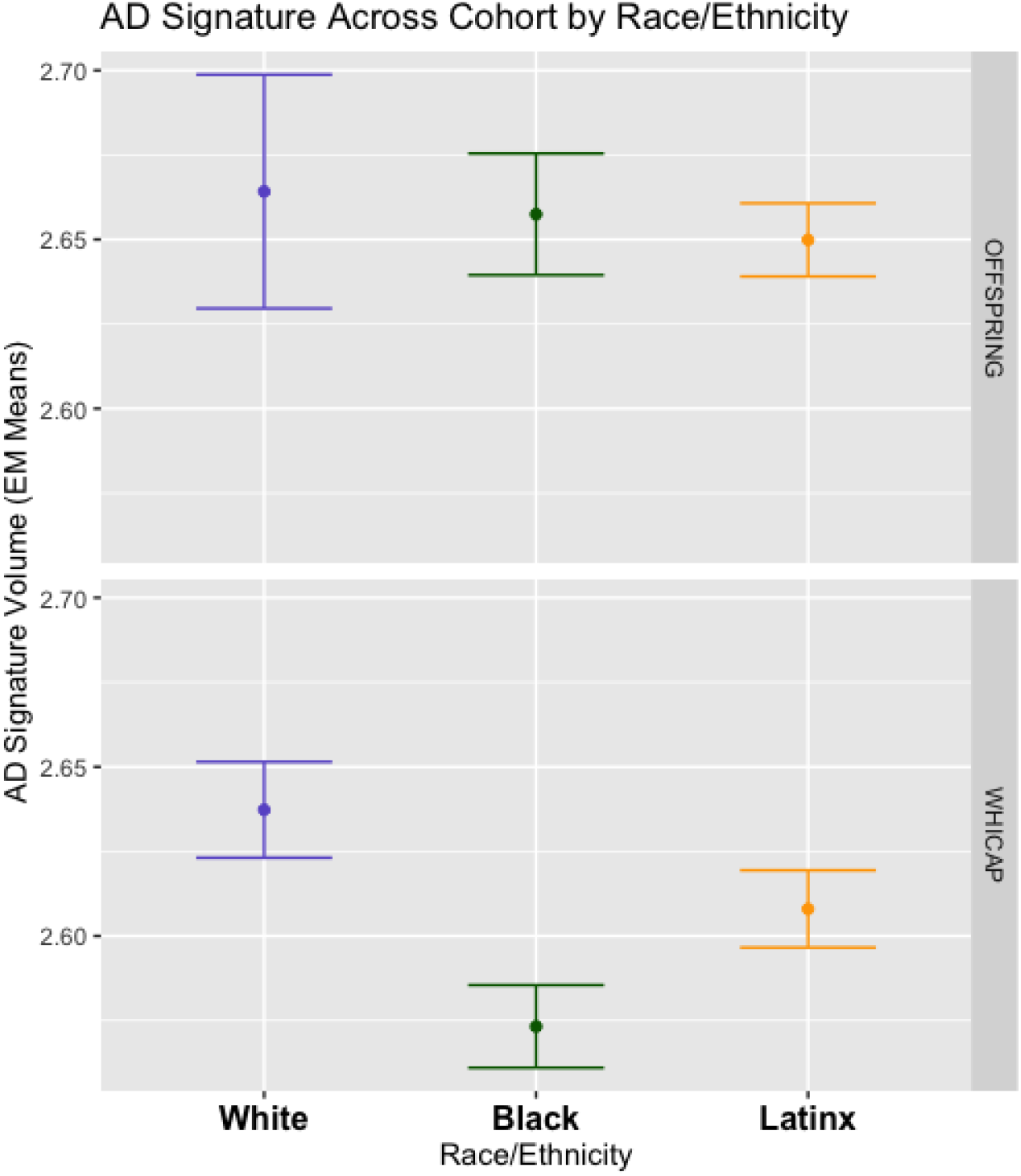
Estimated marginal means of cortical thickness by race and ethnicity across study cohort. Upper panel. In Offspring, white, Black, and Latinx participants had similar cortical thickness. Bottom panel. In WHICAP, Blacks and Latinxs had disproportionately less cortical thickness compared with whites. Latinxs had greater cortical thickness compared to Blacks.

For white matter hyperintensity volumes, in WHICAP, Blacks had greater WMH volume than whites and Latinxs, and Latinxs and whites had comparable WMH volume (main effect of race/ethnicity, F_2,950_=7.07, *p*=.001; η^2^=.02). In Offspring, Blacks had greater WMH volume than whites, but not Latinxs who’s WMH volume was more similar to whites (main effect of race/ethnicity, F_2,413_=3.35, *p*=.036; η^2^=.02). Pairwise effects revealed that the mean difference in WMH volume in Blacks compared with whites were much larger in Offspring than WHICAP (WHICAP Blacks = .103; Offspring Blacks = .209). In both studies, older age was associated with greater WMH volume (Figure 3). Women had lower WMH volume than men in both studies, but this effect was most reliable in Offspring participants. Sensitivity analysis by exclusive middle-age and older age groups resulted in generally the same pattern, with some effects becoming even more robust (not reported).

**Figure 3.**
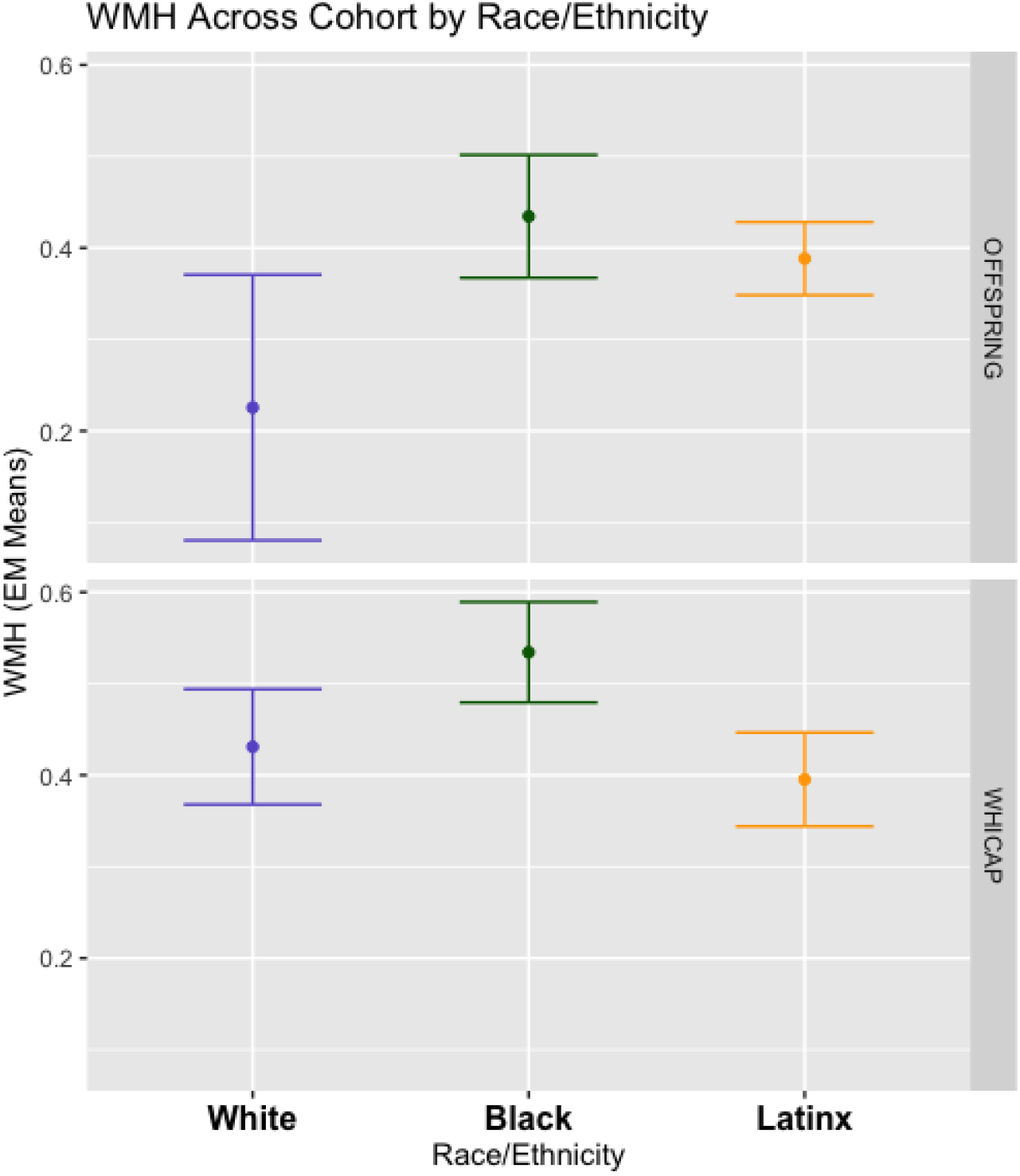
White Matter Hyperintensity Volume by Race and Ethnicity Across Study Cohorts. Upper panel. In Offspring, Blacks, but not Latinxs had greater white matter hyperintensity volume compared with whites. Bottom panel. In WHICAP, Blacks had disproportionately greater white matter hyperintensity volume compared with both Latinxs and whites.

## 4. Discussion

Our study examined the relationship of race and ethnicity on MRI markers of neurodegeneration and cerebrovascular disease among middle-aged and older adults from an urban community. The main finding was that WMH volume differed across race and ethnicity groups within each study (i.e., Offspring and WHICAP). Consistent with previous findings^12^, there were disparities in cortical thickness in late life (i.e., WHICAP study). There was no relationship between race and ethnicity on cortical thickness in midlife (i.e., Offspring study).

Comparable to other reports^13,16^, racial and ethnic differences in WMH burden was observed in later life, where older Blacks had disproportionately greater WMH burden than white and Latinx participants. Most interestingly, this study was the first to show that racial and ethnic differences in WMH volume were already apparent in midlife where Blacks had disproportionately greater WMH burden than whites. Together, our findings show differences in markers of cerebrovascular disease, but not neurodegeneration, among race/ethnicity groups are already apparent in midlife.

Our findings could be explained in part by the weathering hypothesis^10,11^, which provides a framework for interpreting race-related differences in biological processes. Accordingly, the cumulative impact of social, physical, and economic adversities faced by individuals from underrepresented populations lead to early health deterioration and advanced biological aging, which is believed to be caused by chronic or reoccurring stressors. Although not the focus of the “Weathering Hypothesis,” these general ideas might also extend to brain health since research suggests that prolonged exposure to such adversities can have negative impacts on the brain, including increased cerebrovascular disease, resulting in higher WMH volume in Blacks in midlife, compared with whites. Others also suggest that such adversities may reduce the brain’s ability to resist subtle brain damage, increasing the brain’s vulnerability to pathological toxins including AD-related pathology^24,25^.

Further, previous studies reported increased dementia risk factors, cerebrovascular disease, and dementia among Blacks and Latinxs compared with whites^26,27^. We postulate that the racial and ethnic disparities in cortical thickness and especially WMH burden already apparent in midlife are due to lifetime exposure to life stressors, including socioeconomic status (SES) disadvantages and differences in the prevalence of risk factors. Notably, the current study investigated race and ethnicity group differences in MRI markers across middle-aged and older age adults. However, future work will examine the factors that play a role in these associations.

Specifically, environmental (e.g., SES), sociocultural (e.g., institutional racism), behavioral (e.g., discrimination), and biological (e.g., inflammation) factors influence brain integrity and vary tremendously across race and ethnicity groups.

Of importance, when comparing the mean differences between Black and white participants within each cohort, Blacks had much higher WMH volume compared to whites in the Offspring cohort compared with the Black-white difference in WHICAP. This was surprising, especially given that the offspring cohort is over 20 years younger, on average, compared with WHICAP participants. One explanation for the observed narrowing of racial/ethnic disparities in WMH volume is that this effect may be due to a differential survival effect.

Previous findings show that Blacks have higher mortality rates and lower life expectancy^28^, resulting in fewer Blacks surviving to older ages, compared to whites^29,30^. Thus, middle-aged Blacks with disproportionately high WMH volume are less likely to live until older ages, and the survivors in later life exhibit a greater deal of hardiness (i.e., less brain pathology), which would lead to the observed narrowing of racial/ethnic disparities at older ages.

Though many of the associations between MRI markers of neurodegeneration and cerebrovascular disease and race and ethnicity were statistically reliable, effect sizes were small. The magnitude of the relationships suggests that the observed effects are subtle. Thus, future studies will further evaluate their functional values. Future studies should also explore the implications of more severe WMH burden in Blacks compared with whites in midlife.

The WHICAP and Offspring cohorts include participants who are often excluded from studies of brain aging. Data from both studies, drawn from the community, provide a unique opportunity to examine the role of race and ethnicity across a much wider age-range of participants (25-98 years), who also have varied health status and diverse economic and educational backgrounds. Consequently, results from our study more likely reflect the increasing diversity of the general population. This study is the first to compare cortical thickness and WMH burden among middle-aged and older adults across 3 major racial and ethnic groups. Both studies are longitudinal and include comprehensive neuropsychological, medical, and behavioral assessments. Future work will assess the link role of medical risk factors and social determinants of health on these MRI markers in midlife. These assessments will be critical in better understanding of disparity in AD risk across ethnoracial groups.

## Data Availability

Data from the WHICAP or Offspring studies can be requested and will be revised by a committee for approval.

https://cumc.co1.qualtrics.com/jfe/form/SV_6x5rRy14B6vpoqN

## Acknowledgments

Data collection and sharing for this project was supported by the Washington Heights-Inwood Columbia Aging Project (WHICAP: P01AG07232, R01AG03721, RF1AG054023, R56AG034189, R01AG034189, R01AG054520, R01AG028786; Offspring: RF1AG058067, RF1AG054070) funded by the National Institute on Aging (NIA). WHICAP and Offspring investigators have reviewed this manuscript for scientific content and consistency of data interpretation with previous study publications. We acknowledge the WHICAP and Offspring study participants and the research and support staff for their contributions to this study.

## REFERENCES

1. Williams DR. Advancing Our Understanding of Race-related Stressors. Journal of Health and Social Behavior.:20.

2. Williams DR, Mohammed SA. Discrimination and racial disparities in health: evidence and needed research. J Behav Med. 2009;32(1):20–47. doi:10.1007/s10865-008-9185-0

3. Whitfield KE, Thorpe, Jr. RJ. Perspective: Longevity, Stress, Genes and African Americans. Ethn Dis. 27(1):1–2. doi:10.18865/ed.27.1.1

4. National Research Council (US) Panel on Race E and Health in Later Life, Bulatao RA, Anderson NB. Understanding Racial and Ethnic Differences in Health in Late Life: A Research Agenda. Vol 8. National Academies Press (US); 2004. Accessed December 8, 2020. https://www.ncbi.nlm.nih.gov/books/NBK24685/

5. Brewer LC, Redmond N, Slusser JP, et al. Stress and Achievement of Cardiovascular Health Metrics: The American Heart Association Life’s Simple 7 in Blacks of the Jackson Heart Study. J Am Heart Assoc. 2018;7(11). doi:10.1161/JAHA.118.008855

6. Pedersen SS, von Känel R, Tully PJ, Denollet J. Psychosocial perspectives in cardiovascular disease. Eur J Prev Cardiolog. 2017;24(3_suppl):108–115. doi:10.1177/2047487317703827

7. Gruenewald TL, Cohen S, Matthews KA, Tracy R, Seeman TE. Association of socioeconomic status with inflammation markers in Black and White men and women in the coronary artery risk development in young adults (CARDIA) study. Social Science & Medicine. 2009;69(3):451–459. doi:10.1016/j.socscimed.2009.05.018

8. Ranjit N, Diez-Roux AV, Shea S, et al. Psychosocial factors and inflammation in the multi-ethnic study of atherosclerosis. Arch Intern Med. 2007;167(2):174–181. doi:10.1001/archinte.167.2.174

9. Shadlen M-F, Siscovick D, Fitzpatrick AL, Dulberg C, Kuller LH, Jackson S. Education, Cognitive Test Scores, and Black-White Differences in Dementia Risk. Journal of the American Geriatrics Society. 2006;54(6):898–905. doi:https://doi.org/10.1111/j.1532-5415.2006.00747.x

10. Geronimus AT. The weathering hypothesis and the health of African-American women and infants: evidence and speculations. Ethn Dis. 1992;2(3):207–221.

11. Geronimus AT, Pearson JA, Linnenbringer E, et al. Race/Ethnicity, Poverty, Urban Stressors and Telomere Length in a Detroit Community-Based Sample. J Health Soc Behav. 2015;56(2):199–224. doi:10.1177/0022146515582100

12. McDonough IM. Beta-amyloid and Cortical Thickness Reveal Racial Disparities in Preclinical Alzheimer’s Disease. Neuroimage Clin. 2017;16:659–667. doi:10.1016/j.nicl.2017.09.014

13. DeCarli C, Reed BR, Jagust WJ, Martinez O, Ortega M, Mungas D. Brain Behavior Relationships amongst African Americans, Caucasians and Hispanics. Alzheimer Dis Assoc Disord. 2008;22(4):382–391.

14. Brickman AM, Schupf N, Manly JJ, et al. Brain morphology in elderly African Americans, Caribbean Hispanics, and Caucasians from Northern Manhattan. Arch Neurol. 2008;65(8):1053–1061. doi:10.1001/archneur.65.8.1053

15. Manly J, Rentería MA, Avila-Rieger JF, et al. Offspring Study of Racial and Ethnic Disparities in Alzheimer’s Disease: Objectives and Design. PsyArXiv; 2020. doi:10.31234/osf.io/frbkj

16. Brickman AM, Schupf N, Manly JJ, et al. Brain morphology in elderly African Americans, Caribbean Hispanics, and Caucasians from Northern Manhattan. Arch Neurol. 2008;65(8):1053–1061. doi:10.1001/archneur.65.8.1053

17. Brickman AM, Tosto G, Gutierrez J, et al. An MRI measure of degenerative and cerebrovascular pathology in Alzheimer disease. Neurology. 2018;91(15):e1402–e1412. doi:10.1212/WNL.0000000000006310

18. Brickman AM, Muraskin J, Zimmerman ME. Structural neuroimaging in Alzheimer’s disease: do white matter hyperintensities matter? Dialogues Clin Neurosci. 2009;11(2):181–190.

19. Brickman AM, Sneed JR, Provenzano FA, et al. Quantitative approaches for assessment of white matter hyperintensities in elderly populations. Psychiatry Res. 2011;193(2):101–106. doi:10.1016/j.pscychresns.2011.03.007

20. Brickman AM, Provenzano FA, Muraskin J, et al. Regional white matter hyperintensity volume, not hippocampal atrophy, predicts incident Alzheimer’s disease in the community. Arch Neurol. 2012;69(12):1621–1627. doi:10.1001/archneurol.2012.1527

21. Smith SM. Fast robust automated brain extraction. Human Brain Mapping. 2002;17(3):143–155. doi:https://doi.org/10.1002/hbm.10062

22. Isensee F, Schell M, Tursunova I, et al. Automated brain extraction of multi-sequence MRI using artificial neural networks. Hum Brain Mapp. 2019;40(17):4952–4964. doi:10.1002/hbm.24750

23. Tustison NJ, Avants BB, Cook PA, et al. N4ITK: Improved N3 Bias Correction. IEEE Transactions on Medical Imaging. 2010;29(6):1310–1320. doi:10.1109/TMI.2010.2046908

24. Gilbertson MW, Shenton ME, Ciszewski A, et al. Smaller hippocampal volume predicts pathologic vulnerability to psychological trauma. Nature Neuroscience. 2002;5(11):1242–1248. doi:10.1038/nn958

25. Sapolsky RM, Krey LC, McEwen BS. The Neuroendocrinology of Stress and Aging: The Glucocorticoid Cascade Hypothesis*. Endocrine Reviews. 1986;7(3):284–301. doi:10.1210/edrv-7-3-284

26. Tang M-X, Cross P, Andrews H, et al. Incidence of AD in African-Americans, Caribbean Hispanics, and Caucasians in northern Manhattan. Neurology. 2001;56(1):49–56. doi:10.1212/WNL.56.1.49

27. Barnes LL, Bennett DA. Alzheimer’s Disease In African Americans: Risk Factors And Challenges For The Future. Health Aff (Millwood). 2014;33(4):580–586. doi:10.1377/hlthaff.2013.1353

28. Mendes de Leon CF, Barnes LL, Bienias JL, Skarupski KA, Evans DA. Racial Disparities in Disability: Recent Evidence From Self-Reported and Performance-Based Disability Measures in a Population-Based Study of Older Adults. The Journals of Gerontology: Series B. 2005;60(5):S263–S271. doi:10.1093/geronb/60.5.S263

29. Glymour MM, Weuve J, Chen JT. Methodological Challenges in Causal Research on Racial and Ethnic Patterns of Cognitive Trajectories: Measurement, Selection, and Bias. Neuropsychol Rev. 2008;18(3):194–213. doi:10.1007/s11065-008-9066-x

30. Johnson NE. The Racial Crossover in Comorbidity, Disability, and Mortality. Demography. 2000;37(3):267–283. doi:10.2307/2648041

